# A common symptom geometry of mood improvement under sertraline and placebo associated with distinct neural patterns

**DOI:** 10.1101/2023.12.15.23300019

**Authors:** Lucie Berkovitch, Kangjoo Lee, Jie Lisa Ji, Markus Helmer, Masih Rahmati, Jure Demšar, Aleksij Kraljič, Andraž Matkovič, Zailyn Tamayo, John D. Murray, Grega Repovš, John H. Krystal, William J. Martin, Clara Fonteneau, Alan Anticevic

## Abstract

**Importance:** Understanding the mechanisms of major depressive disorder (MDD) improvement is a key challenge to determine effective personalized treatments.

**Objective:** To perform a secondary analysis quantifying neural-to-symptom relationships in MDD as a function of antidepressant treatment.

**Design:** Double blind randomized controlled trial.

**Setting:** Multicenter.

**Participants:** Patients with early onset recurrent depression from the public Establishing Moderators and Biosignatures of Antidepressant Response in Clinical Care (EMBARC) study.

**Interventions:** Either sertraline or placebo during 8 weeks (stage 1), and according to response a second line of treatment for 8 additional weeks (stage 2).

**Main Outcomes and Measures:** To identify a data-driven pattern of symptom variations during these two stages, we performed a Principal Component Analysis (PCA) on the variations of individual items of four clinical scales measuring depression, anxiety, suicidal ideas and manic-like symptoms, resulting in a univariate measure of clinical improvement. We then investigated how initial clinical and neural factors predicted this measure during stage 1. To do so, we extracted resting-state global brain connectivity (GBC) at baseline at the individual level using a whole-brain functional network parcellation. In turn, we computed a linear model for each brain parcel with individual data-driven clinical improvement scores during stage 1 for each group.

**Results:** 192 patients (127 women), age 37.7 years old (standard deviation: 13.5), were included. The first PC (PC1) capturing 20% of clinical variation was similar across treatment groups at stage 1 and stage 2, suggesting a reproducible pattern of symptom improvement. PC1 patients’ scores significantly differed according to treatment during stage 1, whereas no difference of response was evidenced between groups with the Clinical Global Impressions (CGI). Baseline GBC correlated to stage 1 PC1 scores in the sertraline, but not in the placebo group.

**Conclusions and Relevance:** Using data-driven reduction of symptoms scales, we identified a common profile of symptom improvement across placebo and sertraline. However, the neural patterns of baseline that mapped onto symptom improvement distinguished between treatment and placebo. Our results underscore that mapping from data-driven symptom improvement onto neural circuits is vital to detect treatment-responsive neural profiles that may aid in optimal patient selection for future trials.

**Key Points:** *Question:* What is the data-driven pattern of clinical improvement in major depressive disorder across antidepressants and placebo?

*Findings:* Clinical improvement has a shared behavioral geometry across groups but differs in terms of intensity between sertraline and placebo. Baseline resting-state global brain connectivity was correlated to improvement scores in the sertraline group only.

*Meaning:* There is a common behavioral signature of clinical improvement that can be more robustly predicted by baseline neurobehavioral features when it is pharmacologically induced.

## Introduction

Major depressive disorder (MDD) is a devastating and heterogenous psychiatric disease.^1^ The use of a traditional categorical approach, derived from the DSM-5 criteria^2^ severely limits the identification of treatment response predictors.^3^ Clinical trials traditionally use predefined scales to compare treatment to placebo. However, an existing gap in our field relates to whether there could be a generalized response pattern that cuts across active treatment and placebo.^4,5^ Furthermore, it remains unknown if patients’ baseline neural configurations are associated with a unique clinical response pattern. In this study, we intend to test whether a common data driven profile of response can be identified across both treatment and placebo arms. We evaluated data from the publicly available dataset EMBARC (Establishing Moderators and Biosignatures of Antidepressant Response in Clinical Care). EMBARC is a multisite randomized placebo-controlled trial in which unmedicated participants with MDD were treated either by sertraline, a serotonin selective reuptake inhibitor (SSRI), or placebo, and then switched to bupropion (an atypical antidepressant) according to clinical response.^6^ EMBARC collected clinical measures (including item level data on different scales) and baseline neuroimaging data to find markers associated with antidepressant treatment outcomes.^7–13,13–17^ One of the specific aspect of EMBARC is the remarkable intensity of the placebo effect. Using the CGI (Clinical Global Impressions, a clinician rated non-specific 7-point scale providing a single global measure of improvement), or global clinical scores such as the Hamilton Rating Scale for Depression (HRSD), no difference in clinical outcomes was evidenced between the placebo and the sertraline groups.^7,9,10,12,13,18^ However, several predicting factors of sertraline and placebo response were identified, some of them being shared^10,18^ and other being specific to the sertraline or the placebo arm.^7,9,11,13–16^

Here we applied a new analytic strategy to evaluate whether the placebo and the sertraline arms share a common pattern of symptom improvement. Specifically, we performed a Principal Component Analysis (PCA) on the variations of individual items of four clinical scales, resulting in a univariate measure of clinical improvement. We then compare the sertraline and the placebo arms for this measure and investigated how clinical and neural factors at baseline predicted this measure. With this approach, we were able identify a common clinical profile of symptom improvement that occurs across placebo and sertraline. Critically, however, the baseline neural patterns that were related to symptom improvement strongly distinguished between treatment and placebo.

## Material and methods

### Data Collection, Study Design and Clinical Sample

In EMBARC, patients with MDD received sertraline or placebo during 8 weeks. Then, their treatment was adapted according to the CGI for 8 additional weeks^6^ (see Figure 1). Participants underwent a 3T fMRI at baseline and an extensive clinical assessment at 8 and 16 weeks including the Hamilton Rating Scale for Depression (HRSD), the Altman Self-Rating Mania Scale (ASRM), the Concise Health Risk Tracking (CHRT) and the Concise Associated Symptoms Tracking Scale (CASTS). We included 192 patients who had full clinical data and quality-controlled neuroimaging data at baseline (see Figure 1 and Table 1).

**Fig. 1.**
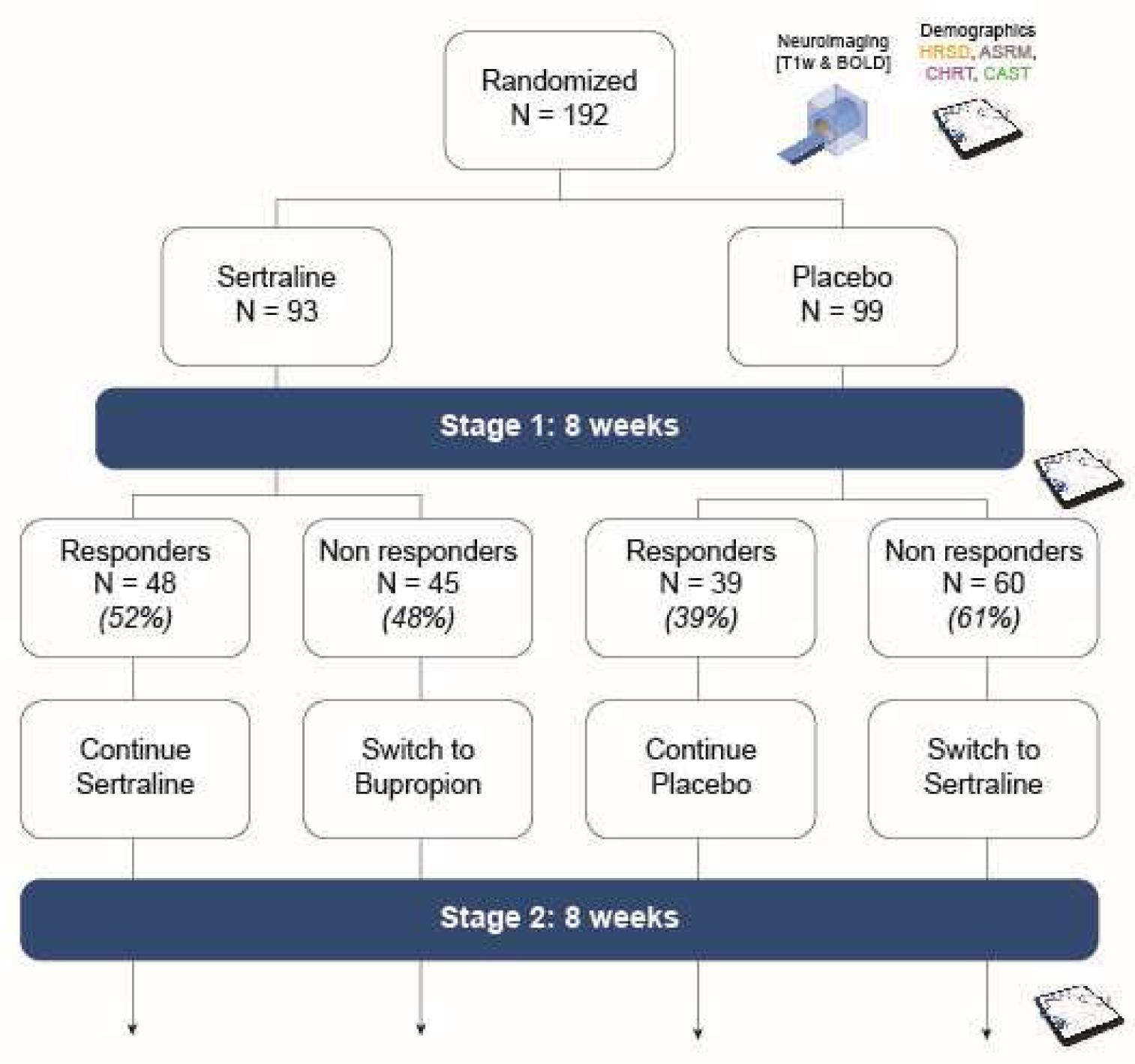
Study design. In Stage 1, patients with early onset recurrent MDD were randomized to receive either sertraline up to 200 mg daily or placebo under double blind conditions. At week 8, participants were assessed with the CGI and those who had a score of less than “much improved” were considered non-responders. In stage 2, non-responding patients in stage 1 were switched to another treatment under double blind conditions: sertraline non-responders received bupropion, and placebo non-responders received sertraline. Responders continued the treatment received during stage 1. Participants underwent an extensive clinical assessment and a 3T fMRI at baseline. At 8 and 16 weeks, four clinical scales of interest were performed: Hamilton Rating Scale for Depression (HRSD) which assesses depressive symptoms, Altman Self-Rating Mania Scale (ASRM) which measures manic symptoms, Concise Health Risk Tracking (CHRT) which evaluates suicidal propensity and risk and Concise Associated Symptoms Tracking Scale (CASTS) which reflects both anxiety, irritability and manic symptoms.

**Table 1.**
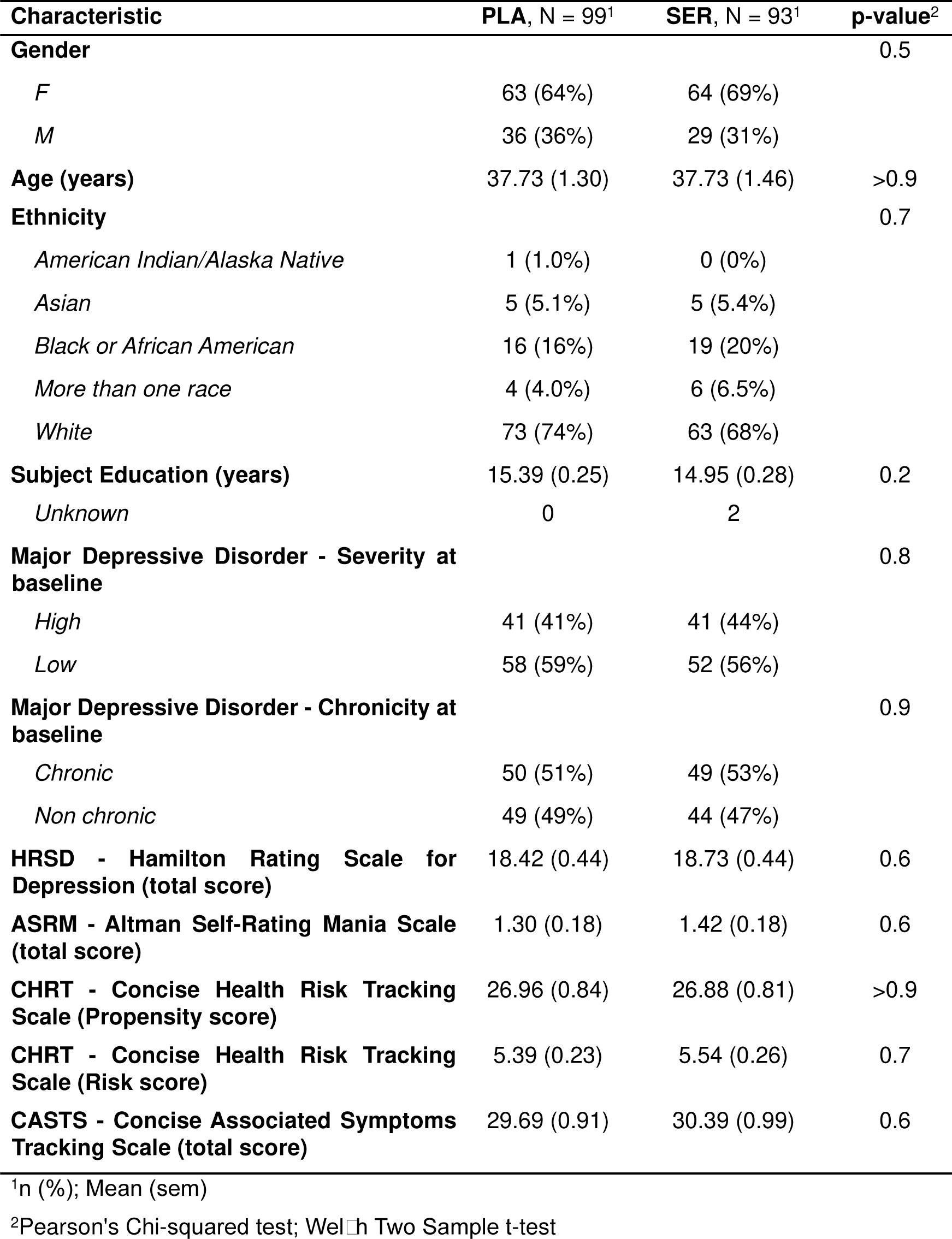
Baseline patients characteristics.

### Dimension reduction of symptom improvement

To estimate a data-driven pattern of symptom improvement, we run a 3-step analysis pipeline on all 73 items across the HRSD, ASRM, CHRT, and CASTS for each individual: (i) dimension reduction of behavioral measures using PCA, separately for each group, (ii) evaluation of PC reproducibility using cross-validation, (iii) comparison of PC loadings across groups to identify shared/distinct axes of symptom improvement. The methodology used for each of these steps is detailed in the Supplementary. PCA can reveal differences between groups in two ways. First, groups can differ in terms of PC geometry (i.e. loadings), meaning that the type of symptoms that change in time depends on the group. Alternatively, groups can have the same PC geometry (i.e. their PC loadings are highly correlated). In such cases, we can rerun the PCA pooling the participants together and show that they have different score distributions for the resulting common PCs (i.e. different “intensity” of change on a common axis of clinical variation), using post-hoc two-sample two-sided t-tests.

### Dimension reduction of neural data

Neuroimaging data were acquired at baseline during resting state using 3T fMRI.^19^ All scans were processed using the Quantitative Neuroimaging Environment & ToolboX (QuNex),^20^ which integrates the Human Connectome Project Pipelines.^21^ The preprocessing pipeline is described in the Supplementary. Neural data was examined at different levels: parcels (718 parcels), networks (12 networks) and whole brain (brain average). Data was first parcellated following the Cole-Anticevic Brain Network Parcellation (CAB-NP) atlas.^22,23^ GBC was then calculated for each subject at the parcel level to reduce the dimensionality of the neural feature space^22^ (see Supplementary about GBC calculation). For network-level analyses, we averaged GBC values across parcels belonging to each network – at the cortical and subcortical levels –, derived from the CAB-NP atlas. For the brain average level analyses, we averaged GBC values across the 718 parcels.

### Mass Univariate Behavioral-to-Neural Mapping

The interactions between behavioral scores and individual baseline GBC at the parcel-level were measured via a mass univariate regression procedure,^22^ through ANOVA with GBC as a dependent variable, behavioral scores during stage 1 (common PC or response status according to the CGI) as an independent variable and age, gender and site as covariates. The resulting maps correspond to the regression coefficients between patients’ behavioral scores and GBC in each parcel, across all 192 patients. This analysis was also run at the network, the subcortical and brain average levels. The greater the magnitude of the coefficient for a given location, the stronger the statistical relationship between GBC and the behavioral variation across patients during stage 1. Significance of the maps was assessed using PALM.^24^ We computed two-tailed Pearson’s correlation, run 1000 permutations and applied family-wise error rate correction.

## Results

### Analysis of global clinical scores

Differences in global clinical scores at baseline and during each stage are detailed in the Supplementary. There was no difference at baseline between treatment groups. The CHRT was the only scale which improved more in the sertraline than in the placebo group during stage 1 (delta of CHRT propensity score: *p* = 0.009; CHRT risk score: *p* = 0.002). The proportion of responders and non-responders according to the CGI was similar in the two groups at the end of stage 1 (placebo 39.4% vs. sertraline 51.6%, *χ*^2^ = 2.4, *p* = 0.12). There were no significant differences at baseline between responders and non-responders (all *p* > 0.08), suggesting that baseline symptoms were not predictive of subsequent clinical response.

### Identifying clinical improvement geometry using Principal Component Analysis

#### The first Principal Component has similar loadings across groups

To further explore the geometry of symptom improvement across groups, we ran separate PCA for each subgroup on the clinical item differences during each stage. Main results are presented in Figure 2 and numeric results are reported in Table 2. Except in patients continuing the same treatment in stage 2, PCA yielded significant and reliable PC1. PC loadings were overall significantly correlated between treatment groups, suggesting that the geometries of symptom improvement were highly similar between groups. To study the distributions of participants’ scores on a common axis of symptom improvement according to their group, we ran an additional PCA on symptom improvement for each stage pooling all the participants together. The common PC1s resulting from this analysis were very similar in terms of loadings to the PC1 observed for each group separately at each stage (see Figure 2, D, I, and Supplementary for a detailed description of items loadings).

**Fig. 2.**
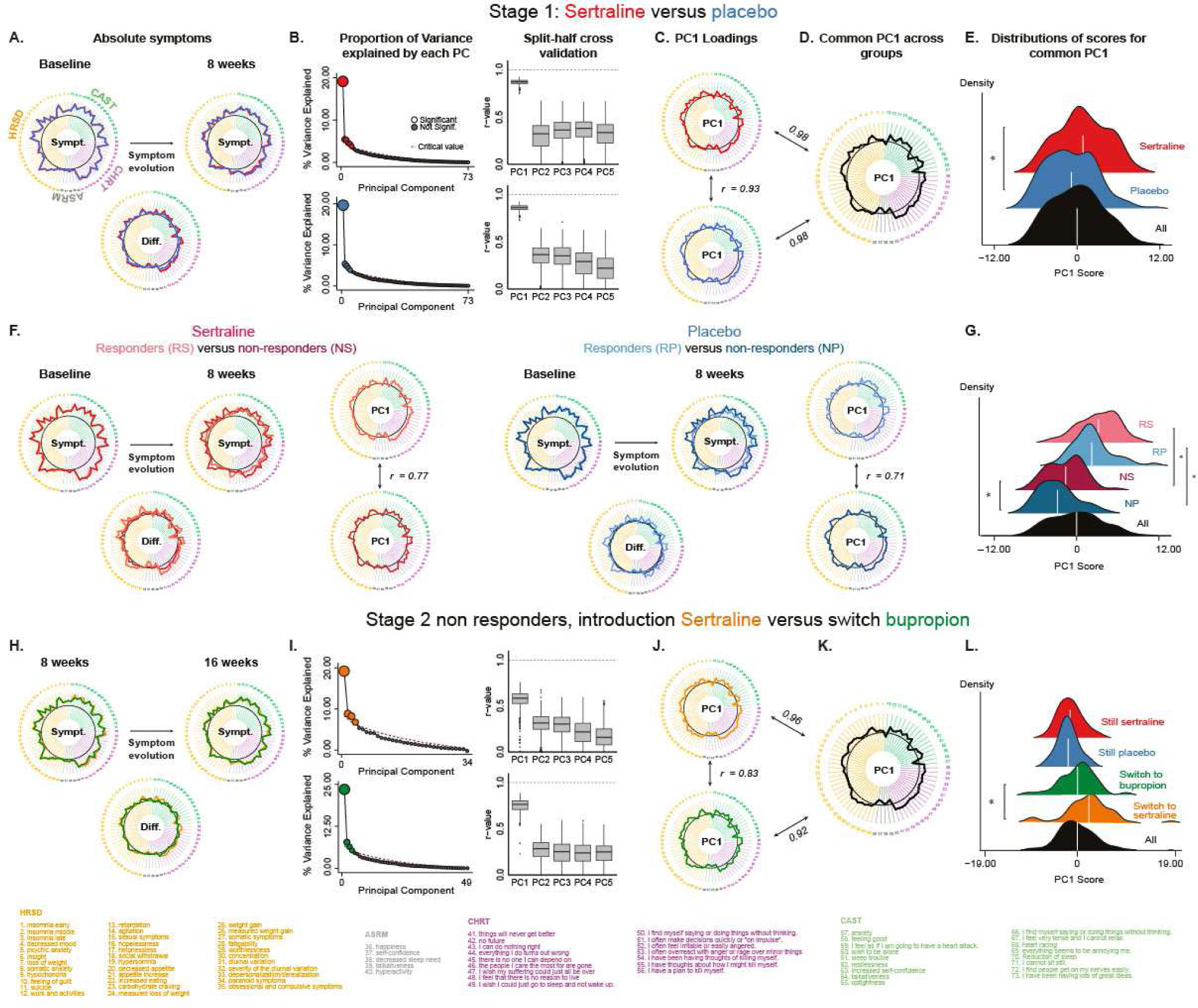
Symptom improvement during stage 1 and 2: absolute evolution and principal components. **A.** Symptom improvement during stage 1 at the item level for each group (red: sertraline, blue: placebo): scores at the item-level are similar between the two treatment groups at baseline and differ only for the CHRT scores evolution. **B.** Proportion of variance explained by PC representing symptom improvement in each group and split-half cross validation (top: sertraline, bottom: placebo). Only PC1, which explains the most variance, is reliable (*r*-value > 0.5). **C.** PC1 loadings: symptom improvement geometry is very similar between the two groups (red: sertraline, blue: placebo). **D.** PCA pooling all participants yields another PC1 (common PC1: black), which is very similar to the PC1 resulting from PCA run separately on the two groups. **E.** Distribution of scores for common PC1 in each group (red: sertraline subgroup, blue: placebo subgroup, black: all participants). On average, patients under sertraline have higher scores than patients under placebo. **F.** Results of the same analyses but splitting each treatment group according to clinical response as measured by CGI (salmon: responders to sertraline, brown: non-responders to sertraline, light blue: responders to placebo, dark blue: non-responders to placebo). Responders and non-responders have significant differences of symptom improvement for all clinical scales. PCA again yields a PC1 that explain most variance and is reliable. **G.** Distribution of scores for the PC1 run across all participants (common PC1) for each subgroup of treatment × response. On average, patients non-responding to sertraline have higher scores than patients non-responding to placebo. **H.** Results of the same analyses performed during stage 2 (green: patients switched from sertraline to bupropion, orange: patients switched from placebo to sertraline). Patients switched to bupropion have lower global CHRT risk score compared to patients switched to sertraline at the beginning of stage 2 and improved less than participants under sertraline during stage 2. **I-J.** PCA again yields a PC1 that explains most variance, is reliable and is relatively similar in terms of loadings between the two groups. **K.** Common PC1 loadings and scores distribution for stage 2 PCA pooling all participants yields another PC1 (common PC1: black), which is very similar to the PC1 resulting from PCA run separately in the different groups. **L.** Distribution of scores for common PC1 in each group for stage 2 (red: responders to sertraline during stage 1, blue: responders to placebo during stage 2, green: switched to bupropion, orange: switched to sertraline, black: all participants). On average, patients who switched to sertraline have higher scores than patients who switched to bupropion.

**Table 2.**
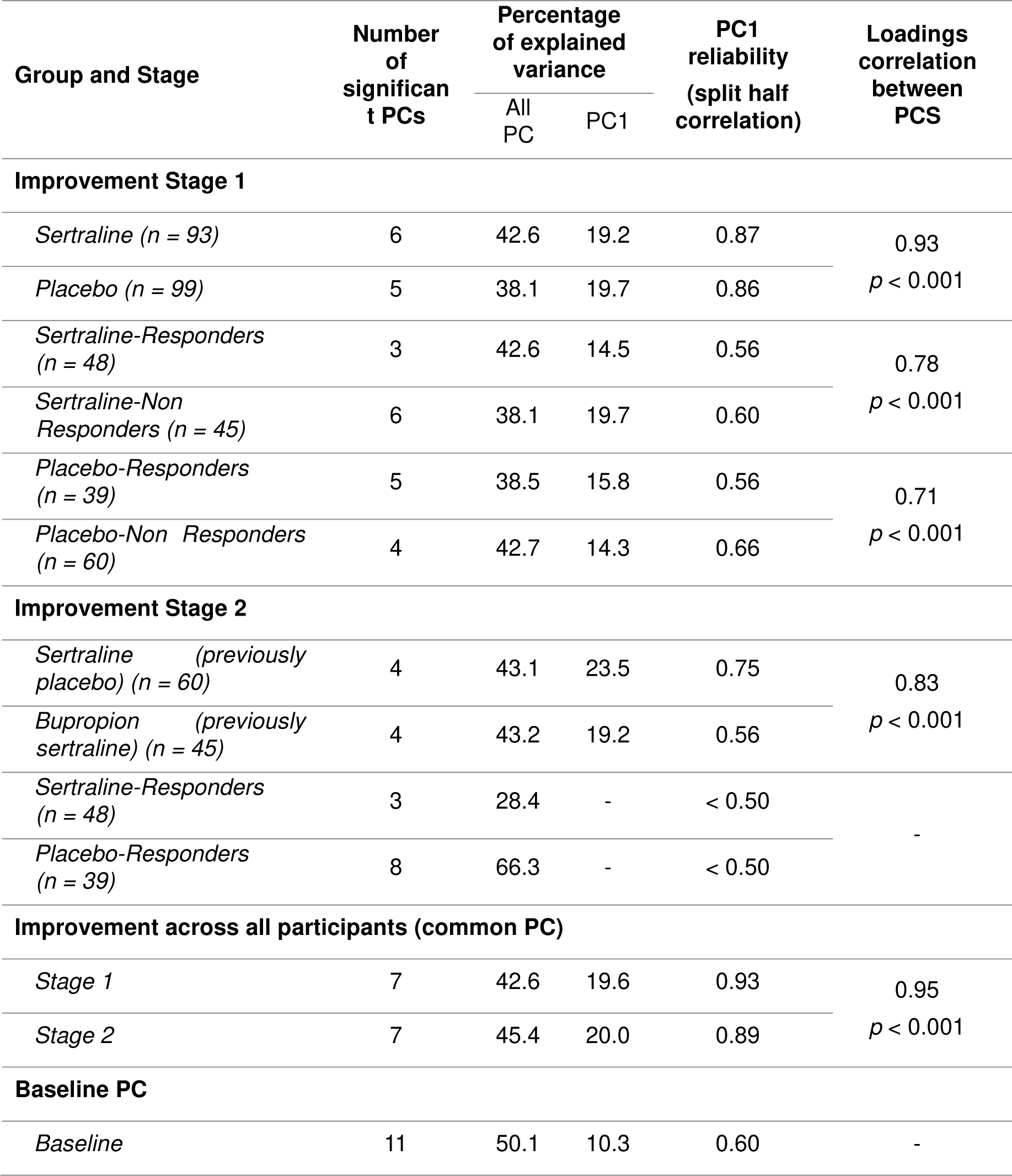
Principal Component Analysis Results.

#### Different PC1 scores distributions between groups

We then explored whether treatment groups had different scores distributions on common PC1. During stage 1, the sertraline group had on average higher common PC1 scores than the placebo group (*t*_190_ = 3.16, *p* = 0.0018, see Figure 2E). Responders also had higher scores than non-responders (sertraline: responders vs. non-responders: *t*_91_ = 8.23, *p* < 0.0001, placebo: responders vs. non-responders: *t*_80_ = 8.57, *p* < 0.0001, Figure 2, panel G), with a significant difference between sertraline versus placebo non-responders (*t*_97_ = 2.25, *p* = 0.027, whereas sertraline versus placebo responders: *t*_82_ = 1.54, *p* = 0.13). During stage 2, the sertraline group had a higher score than the bupropion group (*t*_80_ = 2.39, *p* = 0.019, Figure 2H). However, since those two groups were defined on previous response, those patients were not comparable in terms of treatment-resistance profiles and sample size. The groups that kept stage 1 treatment did not significantly differ in their score distributions (*t*_80_ = 0.82, *p* = 0.42).

#### Baseline predicting factors of clinical improvement

We then explored whether baseline demographic or clinical characteristics could predict PC1 scores during stage 1 by running direct correlation between PC1 scores and baseline clinical scores across subjects and ANOVA with common PC1 scores as a dependent variable and baseline clinical scores and treatment group as independent variables. CHRT risk and CAST global scores had a significant main effect on PC1 scores across groups (CHRT: r = 0.20, *p* = 0.007; CAST: r = 0.20, *p* = 0.006) with no significant differences between the two groups (interaction CHRT × treatment: *F*_1,188_ = 0.6, *p* = 0.45; interaction CAST × treatment: *F*_1,188_ = 0.4, *p* = 0.52). By contrast HRSD scores and depression severity did not have a main effect on PC1 scores but impacted differently PC1 between the sertraline and the placebo group (interaction HRSD × treatment: *F*_1,188_ = 5.8, *p* = 0.017; interaction depression severity × treatment: *F*_1,188_ = 5.6, *p* = 0.019). They had a higher influence on clinical outcome in the sertraline group (HRSD sertraline group: r = 0.21, *p* = 0.040 vs. placebo group: r = −0.13, *p* = 0.19; depression severity (high > low) sertraline group: *F*_1,91_ = 3.7, *p* = 0.058 vs. placebo group: *F*_1,91_ = 2.1, *p* = 0.15). So far, we used PCA to explore clinical improvement, with the delta of the symptoms as inputs. However, PCA can also be run on baseline symptoms to explore whether baseline PCs predict subsequent improvement. Interestingly, the loadings of baseline PC1 and of common improvement PC1 were significantly correlated (r = 0.69, *p* < 0.001), indicating that the improvement mainly concerns symptoms that are observable at baseline. Distribution scores for baseline PC1 did not significantly differ between the sertraline and the placebo groups (*t*_186_ = - 0.82, *p* = 0.41) but significantly correlated with improvement PC1 scores distribution across groups (r = 0.20, *p* = 0.005) with no significant differences between the two groups (interaction baseline PC1 scores × treatment: *F*_1,188_ = 0.001, *p* = 0.98). By contrast, clinical response measured by CGI was not significantly predicted by any baseline characteristics (age, gender, ethnicity, education, MDD severity, MDD chronicity, HRSD, ASMR, CHRT, CAST, baseline PC1, all *p* > 0.08 across subjects and within each treatment group).

### Brain-behavior mapping

In order to identify neural predicting factors of response, we then explored the relationships between the baseline parcellated resting-state GBC and clinical improvement. Importantly, we did not find any differences between the sertraline and the placebo groups in baseline GBC (with age, gender and site as covariates) at the parcel, network, and brain average levels (all *p_adjusted_* > 0.1).

#### Parcellated resting-state GBC

At the parcel level (n = 718), the interactions between GBC and common PC1 scores of improvement, on the one hand, or the response status (CGI), on the other hand, did not survive correction for multiple comparisons (family-wise error rate correction, *α*=0.05 in PALM^25^). The corresponding neurobehavioral correlation maps are shown in Figure 3A. Additional exploratory analyses revealed a higher correlation between the GBC-PC1 neurobehavioral map and the GBC-response neurobehavioral map in the placebo compared to the sertraline group.

**Fig. 3.**
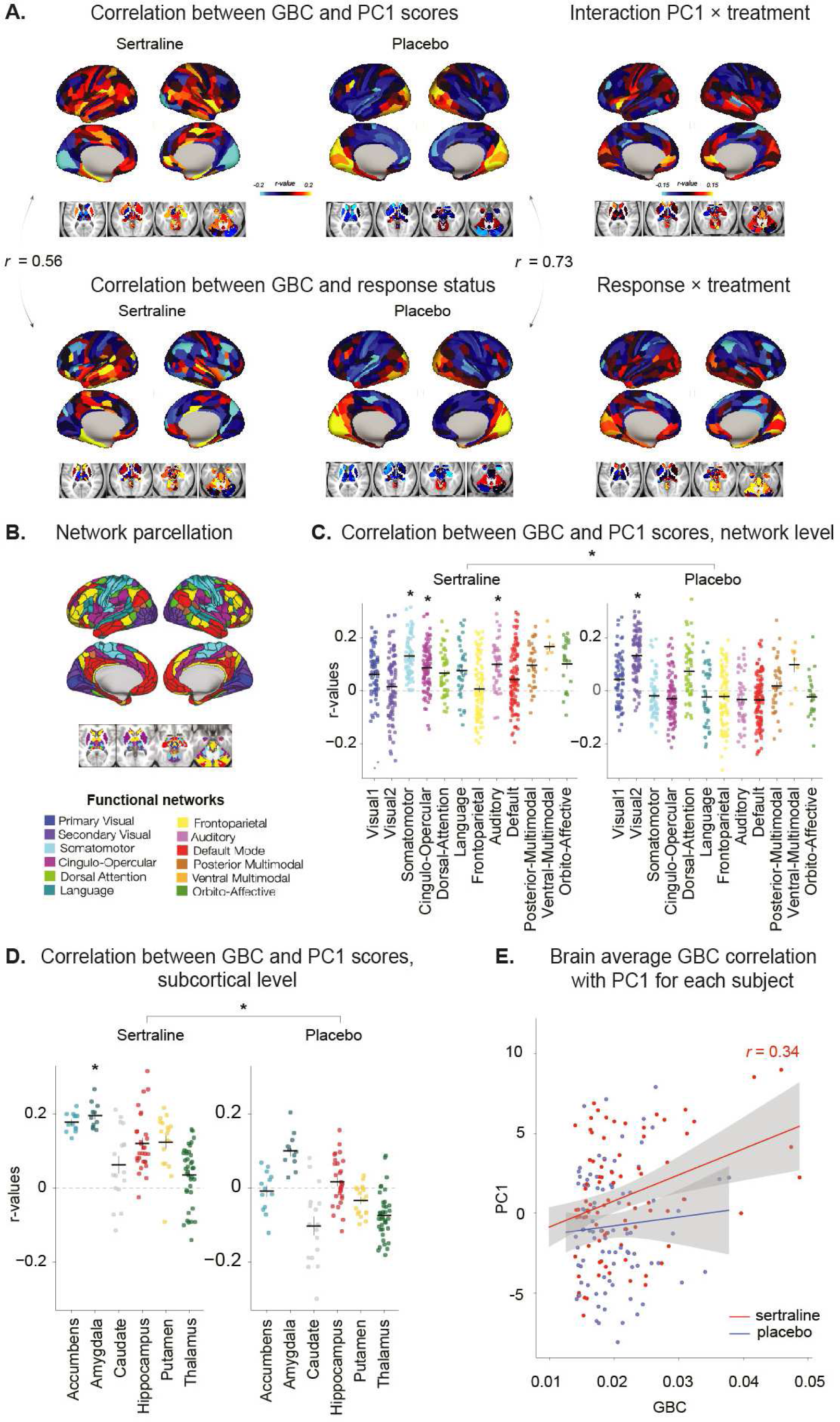
Neurobehavioral mapping of mood improvement during stage 1. **A.** Top. Correlation between the parcellated resting-state GBC and the PC1 scores in the sertraline (left) and the placebo (middle) groups during stage 1, and interaction between the two treatment groups (right). Bottom. Correlation between the parcellated resting-state GBC and the response status in the sertraline (left) and the placebo (middle) groups, and interaction between the two treatment groups (right). Correlation maps are visually different between the sertraline and the placebo groups, suggesting that the baseline cerebral predictors of clinical improvement differ according to the pharmacological intervention. Additional exploratory analyses showed a lower correlation between GBC-PC1 neurobehavioral mapping and GBC-response neurobehavioral map in the sertraline group (r = 0.56, *p* < 0.001) compared to the placebo group (r = 0.73, *p* < 0.001), suggesting that response according to CGI has the same brain predictive factors as PC1 for placebo but not for sertraline. The interaction between GBC, PC1 and treatment on the one hand, and GBC, response and treatment on the other hand, displayed in the right panel show how each parcel contributes to the pharmacological response (as opposed to the placebo effect). It should be noted that those interactions did not reach significance after family-wise error rate correction. **B.** Network parcellation. **C.** Correlation between the parcellated resting-state GBC regrouped by networks and the PC1 scores in the sertraline (left) and the placebo (right) groups. Each dots represents a parcel and each horizontal bar represents the mean of correlation *r* values for a given network across subjects, asterisks indicate significant *p* values (under 0.05), obtained through an ANOVA including also age, gender and site, after Bonferroni correction for multiple comparisons, error bars represent the standard error of the means. There was a significant interaction between PC1 scores and GBC in the sertraline group for the auditory, the cingulo-opercular, and the somatomotor networks, and in the placebo group for one of the two visual network (secondary visual network, Visual 2). *R* values were significantly higher in the sertraline than in the placebo group when compared with a t-test between the two groups. **D.** Correlation between the parcellated resting-state GBC regrouped by subcortical regions and the PC1 scores in the sertraline (left) and the placebo (right) groups. Each dots represents a parcel and each horizontal bar represents the mean of correlation *r* values for a given subcortical region across subjects, asterisks indicate significant *p* values (under 0.05), obtained through an ANOVA including also age, gender and site, after Bonferroni correction for multiple comparisons, error bars represent the standard error of the means. The only subcortical region in which GBC significantly interacted with PC1 score was the amygdala in the sertraline group. *R*-values were significantly higher in the sertraline than in the placebo group when compared with a t-test between the two groups. **E.** Brain average GBC correlation with PC1 in the two treatment groups during stage 1 (red: sertraline, blue: placebo). Each dot represent a subject, lines represent the linear regressions, and the shaded areas represent the 95% confidence interval. PC1 scores and GBC being significantly correlated in the sertraline group, but not in the placebo group, indicating that baseline GBC is a predictive factor of pharmacologically-induced clinical improvement.

#### Network resting-state GBC

At the network level (averaged GBC across parcels belonging to each network, n = 12 as a dependent variable, PC1 as an independent variable, with age, gender and site as covariates, Figure 3C), there was a significant effect of PC1 scores on GBC in the sertraline group for the auditory (*F*_1,86_ = 10.03, *p_adjusted_* = 0.026), the cingulo-opercular (*F*_1,86_ = 8.77, *p_adjusted_* = 0.045), and the somatomotor networks (*F*_1,86_ = 10.17, *p_adjusted_* = 0.024). In the placebo group, only the secondary visual network GBC significantly interacted with PC1 scores after correction (*F*_1,92_ = 13.36, *p_adjusted_* = 0.005). *R*-values were significantly higher in the sertraline than in the placebo group (paired t-test: mean of the difference: 0.06; *t*_11_ = 2.97, *p* = 0.013) with a significant interaction between PC1 scores and treatment for the auditory (*F*_1,183_ = 8.84, *p_adjusted_* = 0.040) and the cingulo-opercular networks (*F*_1,183_ = 9.02 *p_adjusted_* = 0.036), after Bonferroni correction.

#### Subcortical resting-state GBC

At the subcortical level (averaged GBC across parcels belonging to subcortical structure of interest, i.e. amygdala, hippocampus, thalamus and striatum with its substructures: nucleus accumbens, putamen and caudate nucleus, n = 4, with age, gender and site as covariates, Figure 3D), the only subcortical structure in which GBC significantly interacted with PC1 score was the amygdala in the sertraline group (*F*_1,86_ = 7.08, *p_adjusted_* = 0.037). *R*-values were significantly higher in the sertraline than in the placebo group (paired t-test: mean of the difference: 0.10; *t*_10_ = 6.70, *p* < 0.001) with a significant interaction between PC1 scores and treatment for all subcortical regions (all *p_adjusted_* < 0.008, after Bonferroni correction).

#### Brain average resting-state GBC

Brain average resting-state GBC (averaged across 718 parcels, with age, gender and site as covariates) significantly predicted clinical improvement in the sertraline group. First, we explore differences of GBC (dependent variable) according to response status (CGI) or PC1 scores (independent variables). GBC was higher for responders compared to non-responders in the sertraline group only (interaction response × treatment across participants: *F*_1,183_ = 5.24, *p* = 0.023; sertraline group: *F*_1,86_ = 7.91, *p* = 0.006, placebo: *F*_1,92_ = 0.27, *p* = 0.60) and was significantly correlated with PC1 scores in the sertraline group only (r = 0.34, *t*_91_ = 3.41, *p* = 0.0010; placebo: r = 0.08, *t*_97_ = 0.75, *p* = 0.46). Second, there was a significant effect of GBC on PC1 scores in the sertraline group only (ANOVA with PC1 scores as a dependent variable and GBC, age, gender and site as independent variables: sertraline: *F*_1,86_ = 11.42, *p* = 0.0011, placebo *F*_1,92_ = 0.59, *p* = 0.44, Figure 3E), indicating that baseline GBC is predictive of clinical response to pharmacological intervention but not to placebo.

## Discussion

We studied patterns of mood improvement in a cohort of patients with MDD from the EMBARC clinical trial, treated either with antidepressant (sertraline or bupropion) or placebo, using PCA across multiple clinical scales. This data-driven approach yielded a single dimension (PC1) that passed split-half cross-validation testing. This robust PC solution provided a low dimensional pattern of mood improvement and was strikingly similar across both treatment and placebo, suggesting that mood improvement may evolve along a common symptom axis.

One of the specific aspect of EMBARC is the intensity of the reported placebo response. Previous examinations of EMBARC found no differences between sertraline and placebo efficacy,^7,9,10,12,13,18^ and commonalities in predictive factors between the two groups,^8,10,12,13,18^ supporting the idea that a placebo response is embedded into the antidepressant response.^26^ In our analysis, no difference between the two groups and no clinical predictive factors could be isolated using the CGI only. However, by computing a PCA solution, we show a higher efficacy in the sertraline group that was associated with both behavioral and neural predictive factors. Critically, we included a wide range of items across multiple scales, which increased the sensitivity of the PCA-derived measure^27–29^ (see Supplementary for additional discussion). Indeed, suicidal risk and anxiety scores account for a significant part of symptom variation in the PCA solution, especially in the sertraline arm. In this sense, the efficacy of sertraline above placebo would mostly consist in an “antisuicidal” and an “anxiolytic” effect, whereas the pure “antidepressant” response may be attributed to a placebo effect. Interestingly, we observed a dissociation between predictive factors and improvement dimensions. In line with previous studies, HRSD predicts sertraline efficacy as measured by PC1,^7,30,31^ even if HRSD scores were not more improved in the sertraline compared to the placebo group.^7,9,10,12,13,18^

In turn, we quantified baseline patterns of data-driven functional connectivity and tested if there may be distinct patterns of baseline GBC that would uniquely map onto magnitude of PC-derived symptom improvement in each arm. Indeed we found that PC1 scores were predicted by an increased resting-state GBC at baseline in the sertraline group, notably in the parcels corresponding to the amygdala and to several cerebral networks. Those results are compatible with previous explorations of EMBARC using different approaches,^11,15,32^ and previous studies on independent datasets.^33–41^ However, contrary to previous findings, there was no significant predictive effect of the DMN^9,11,15,42,43^ or of the rostral anterior cingulate cortex connectivity^13,18,44–48^ on clinical improvement. Similarly, previous work shows that a decreased amygdala connectivity at baseline correlates with positive clinical outcomes,^40,49–52^ whereas we, and others,^53^ found an opposite pattern. Those mixed results may be due to the use of GBC for each parcel rather than predetermined ROI or seed, or connectivity measures (within or intrinsic) restricted to networks. More importantly, and in line with previous results,^8,9,11,13,15^ our study shows that baseline neural patterns that relate to sertraline and placebo effects are in fact different. In other words, even if the pattern of the mood improvement is highly similar in the two groups, it seems to be related to very different baseline neural system configuration. The parcel-level analyses further suggest that improvement as measured by CGI and PC1 correspond to distinct neural circuits in the sertraline group but not in the placebo group. Sertraline could therefore activate additional brain mechanisms that may complement or amplify the placebo response (see Supplementary for additional discussion).

Our study has several limitations. First, it is a secondary analysis of a publicly available dataset. This analysis was therefore not initially planned and could have been biased by previously published data on the same cohort. Second, we did not reproduce all previously published findings, probably because we used a different analytical approach and selected a subset of participants who had clinical measures at different time-points, which raises the question of generalizability. In this perspective, our results would highly benefit from a replication with an independent dataset, notably to explore whether the pattern of mood improvement that we observed is specific to this population or more universal. Third, the design did not include neuroimaging at the end of stage 1, so we could not map clinical changes to neural evolution and could only study predictive factors of clinical response. A longitudinal study would allow measuring how neural patterns evolve with time according to treatment and response. Finally, only few conclusions could be drawn from the second stage since participants were assigned to a new treatment according to their clinical response in stage 1, rendering the groups incomparable. Comparing different drugs in randomized parallel arms would allow examining response and predictive factors of various antidepressants, and better characterize their pharmacological mechanisms of action.

In summary, we discovered a common behavioral signature of clinical improvement along the mood spectrum with multiple symptomatic dimensions, on which patients score differently according to the treatment received. At the behavioral level, improvement under antidepressant therefore corresponds to an amplification of the placebo response. This improvement was more robustly predicted by baseline GBC (at different levels) and clinical features in the sertraline group, suggesting that pharmacological improvement relies on more reproducible and specific neurobehavioral features.

## Data Availability

All data produced in the present study are available upon reasonable request to the authors

## Declarations

A.A. and J.D.M. hold equity with Neumora Therapeutics (formerly BlackThorn Therapeutics), Manifest Technologies, and are co-inventors on the following patents: Anticevic A, Murray JD, Ji JL: Systems and Methods for Neuro-Behavioral Relationships in Dimensional Geometric Embedding(N-BRIDGE), PCT International Application No.PCT/US2119/022110, filed March 13, 2019 and Murray JD, Anticevic A, Martin WJ: Methods and tools for detecting, diagnosing, predicting, prognosticating, or treating a neurobehavioral phenotype in a subject, U.S. Application No.16/149,903, filed on October 2, 664 2018, U.S. Application for PCT International Application No.18/054, 009 filed on October 2, 2018. J.L.J. is an employee of Manifest Technologies, has previously worked for Neumora, and is a co-inventor on the following patent: Anticevic A, Murray JD, Ji JL: Systems and Methods for Neuro-Behavioral Relationships in Dimensional Geometric Embedding (N-BRIDGE), PCT International Application No.PCT/US2119/022110, filed March 13, 2019. C.F. consults for Manifest Technologies and formerly consulted for RBNC (formerly BlackThorn Therapeutics). Z.T. has previously consulted for Neumora and consults for Manifest Technologies. G.R. holds equity and consults in Neumora and Manifest Technologies. J.D. and A.K. consult for Neurotherapeutix Medical Services. A.K. and A.M. have previously consulted for Neumora. J.H.K. holds equity in Biohaven Pharmaceuticals, Biohaven Pharmaceuticals Medical Sciences, Clearmind Medicine, EpiVario, Neumora Therapeutics, Tempero Bio, Terran Biosciences, Tetricus, and Spring Care. J.H.K. consults for AE Research Foundation, Aptinyx, Biohaven Pharmaceuticals, Biogen, Bionomics, Limited (Australia), BioXcel Therapeutics, Boehringer Ingelheim International, Cerevel Therapeutics, Clearmind Medicine, Cybin IRL, Delix Therapeutics, Eisai, Enveric Biosciences, Epiodyne, EpiVario, Evidera, Freedom Biosciences, Janssen Research & Development, Jazz Pharmaceuticals, Leal Therapeutics, Neumora Therapeutics, Neurocrine Biosciences, Novartis Pharmaceuticals Corporation, Otsuka America Pharmaceutical, Perception Neuroscience, Praxis Precision Medicines, PsychoGenics, Spring Care, Sunovion Pharmaceuticals, Takeda Industries, Tempero Bio, Terran Biosciences, and Tetricus. All other co-authors declare no competing interests.

## Acknowledgements

L.B. was supported by the Fondation Bettencourt Schueller, the Philippe Foundation and the L’Oréal-Unesco Foundation. G.R., J.D. are funded by ARIS grants P3-0338 and J5-4590, A.K. and A.M. by ARIS grants J5-4590 and P5-0110, K.L. is funded by 1U01MH124639-03 (NIMH/NIH/DHHS); Boehringer Ingelheim International GmbH - Project No.668012.

## References

1. World Health Organization. Depression and Other Common Mental Disorders: Global Health Estimates. World Health Organization; 2017. Accessed August 13, 2021. https://apps.who.int/iris/handle/10665/254610

2. American Psychiatric Association. Diagnostic and Statistical Manual of Mental Disorders: DSM-5. 5th edition. American Psychiatric Association; 2013. Accessed April 28, 2023. https://dsm.psychiatryonline.org/doi/book/10.1176/appi.books.9780890425787

3. Perlman K, Benrimoh D, Israel S, et al. A systematic meta-review of predictors of antidepressant treatment outcome in major depressive disorder. J Affect Disord. 2019;243:503–515. doi:10.1016/j.jad.2018.09.067

4. Gueorguieva R, Mallinckrodt C, Krystal JH. Trajectories of depression severity in clinical trials of duloxetine: insights into antidepressant and placebo responses. Arch Gen Psychiatry. 2011;68(12):1227–1237. doi:10.1001/archgenpsychiatry.2011.132

5. Huneke NTM, Aslan IH, Fagan H, et al. Functional Neuroimaging Correlates of Placebo Response in Patients With Depressive or Anxiety Disorders: A Systematic Review. Int J Neuropsychopharmacol. 2022;25(6):433–447. doi:10.1093/ijnp/pyac009

6. Trivedi MH, McGrath PJ, Fava M, et al. Establishing moderators and biosignatures of antidepressant response in clinical care (EMBARC): Rationale and design. J Psychiatr Res. 2016;78:11–23. doi:10.1016/j.jpsychires.2016.03.001

7. Webb CA, Trivedi MH, Cohen ZD, et al. Personalized prediction of antidepressant v. placebo response: evidence from the EMBARC study. Psychol Med. 2019;49(7):1118–1127. doi:10.1017/S0033291718001708

8. Zhao K, Xie H, Fonzo GA, et al. Individualized fMRI connectivity defines signatures of antidepressant and placebo responses in major depression. Mol Psychiatry. Published online February 2, 2023:1–10. doi:10.1038/s41380-023-01958-8

9. Chin Fatt CR, Jha MK, Cooper CM, et al. Effect of Intrinsic Patterns of Functional Brain Connectivity in Moderating Antidepressant Treatment Response in Major Depression. Am J Psychiatry. 2020;177(2):143–154. doi:10.1176/appi.ajp.2019.18070870

10. Fan S, Nemati S, Akiki TJ, et al. Pretreatment Brain Connectome Fingerprint Predicts Treatment Response in Major Depressive Disorder. Chronic Stress. 2020;4:2470547020984726. doi:10.1177/2470547020984726

11. Chin Fatt CR, Minhajuddin A, Jha MK, Mayes T, Rush AJ, Trivedi MH. Data driven clusters derived from resting state functional connectivity: Findings from the EMBARC study. J Psychiatr Res. 2023;158:150–156. doi:10.1016/j.jpsychires.2022.12.002

12. Whitton AE, Webb CA, Dillon DG, et al. Pretreatment Rostral Anterior Cingulate Cortex Connectivity With Salience Network Predicts Depression Recovery: Findings From the EMBARC Randomized Clinical Trial. Biol Psychiatry. 2019;85(10):872–880. doi:10.1016/j.biopsych.2018.12.007

13. Cooper CM, Chin Fatt CR, Jha M, et al. Cerebral Blood Perfusion Predicts Response to Sertraline versus Placebo for Major Depressive Disorder in the EMBARC Trial. EClinicalMedicine. 2019;10:32–41. doi:10.1016/j.eclinm.2019.04.007

14. Chin Fatt CR, Cooper CM, Jha MK, et al. Differential response to SSRI versus Placebo and distinct neural signatures among data-driven subgroups of patients with major depressive disorder. J Affect Disord. 2021;282:602–610. doi:10.1016/j.jad.2020.12.102

15. Chin Fatt CR, Cooper C, Jha MK, et al. Dorsolateral Prefrontal Cortex and Subcallosal Cingulate Connectivity Show Preferential Antidepressant Response in Major Depressive Disorder. Biol Psychiatry Cogn Neurosci Neuroimaging. 2021;6(1):20–28. doi:10.1016/j.bpsc.2020.06.019

16. Ang YS, Bruder GE, Keilp JG, et al. Exploration of baseline and early changes in neurocognitive characteristics as predictors of treatment response to bupropion, sertraline, and placebo in the EMBARC clinical trial. Psychol Med. 2022;52(13):2441–2449. doi:10.1017/S0033291720004286

17. Beliveau V, Hedeboe E, Fisher PM, et al. Generalizability of treatment outcome prediction in major depressive disorder using structural MRI: A NeuroPharm study. NeuroImage Clin. 2022;36:103224. doi:10.1016/j.nicl.2022.103224

18. Pizzagalli DA, Webb CA, Dillon DG, et al. Pretreatment Rostral Anterior Cingulate Cortex Theta Activity in Relation to Symptom Improvement in Depression: A Randomized Clinical Trial. JAMA Psychiatry. 2018;75(6):547–554. doi:10.1001/jamapsychiatry.2018.0252

19. Greenberg T, Chase HW, Almeida JR, et al. Moderation of the Relationship Between Reward Expectancy and Prediction Error-Related Ventral Striatal Reactivity by Anhedonia in Unmedicated Major Depressive Disorder: Findings From the EMBARC Study. Am J Psychiatry. 2015;172(9):881–891. doi:10.1176/appi.ajp.2015.14050594

20. Ji JL, Demšar J, Fonteneau C, et al. QuNex—An integrative platform for reproducible neuroimaging analytics. Front Neuroinformatics. 2023;17. Accessed October 29, 2023. https://www.frontiersin.org/articles/10.3389/fninf.2023.1104508

21. Glasser MF, Sotiropoulos SN, Wilson JA, et al. The minimal preprocessing pipelines for the Human Connectome Project. NeuroImage. 2013;80:105–124. doi:10.1016/j.neuroimage.2013.04.127

22. Ji JL, Spronk M, Kulkarni K, Repovš G, Anticevic A, Cole MW. Mapping the human brain’s cortical-subcortical functional network organization. NeuroImage. 2019;185:35–57. doi:10.1016/j.neuroimage.2018.10.006

23. Glasser MF, Coalson TS, Robinson EC, et al. A multi-modal parcellation of human cerebral cortex. Nature. 2016;536(7615):171–178. doi:10.1038/nature18933

24. Winkler AM, Ridgway GR, Webster MA, Smith SM, Nichols TE. Permutation inference for the general linear model. NeuroImage. 2014;92(100):381–397. doi:10.1016/j.neuroimage.2014.01.060

25. Smith SM, Nichols TE. Threshold-free cluster enhancement: addressing problems of smoothing, threshold dependence and localisation in cluster inference. NeuroImage. 2009;44(1):83–98. doi:10.1016/j.neuroimage.2008.03.061

26. Peciña M, Bohnert ASB, Sikora M, et al. Association Between Placebo-Activated Neural Systems and Antidepressant Responses: Neurochemistry of Placebo Effects in Major Depression. JAMA Psychiatry. 2015;72(11):1087–1094. doi:10.1001/jamapsychiatry.2015.1335

27. Maier W, Philipp M. Improving the Assessment of Severity of Depressive States: A Reduction of the Hamilton Depression Scale. Pharmacopsychiatry. 1985;18(01):114–115. doi:10.1055/s-2007-1017335

28. Möller HJ. Methodological aspects in the assessment of severity of depression by the Hamilton Depression Scale. Eur Arch Psychiatry Clin Neurosci. 2001;251 Suppl 2:II13–20. doi:10.1007/BF03035121

29. Demyttenaere K, De Fruyt J. Getting What You Ask For: On the Selectivity of Depression Rating Scales. Psychother Psychosom. 2003;72(2):61–70. doi:10.1159/000068690

30. Dodd S, Berk M. Predictors of antidepressant response: A selective review. Int J Psychiatry Clin Pract. 2004;8(2):91–100. doi:10.1080/13651500410005423

31. De Carlo V, Calati R, Serretti A. Socio-demographic and clinical predictors of non-response/non-remission in treatment resistant depressed patients: A systematic review. Psychiatry Res. 2016;240:421–430. doi:10.1016/j.psychres.2016.04.034

32. Rolle CE, Fonzo GA, Wu W, et al. Cortical Connectivity Moderators of Antidepressant vs Placebo Treatment Response in Major Depressive Disorder: Secondary Analysis of a Randomized Clinical Trial. JAMA Psychiatry. 2020;77(4):397–408. doi:10.1001/jamapsychiatry.2019.3867

33. Korgaonkar MS, Goldstein-Piekarski AN, Fornito A, Williams LM. Intrinsic connectomes are a predictive biomarker of remission in major depressive disorder. Mol Psychiatry. 2020;25(7):1537–1549. doi:10.1038/s41380-019-0574-2

34. Strege MV, Siegle GJ, Richey JA, Krawczak RA, Young K. Cingulate prediction of response to antidepressant and cognitive behavioral therapies for depression: Meta-analysis and empirical application. Brain Imaging Behav. Published online January 9, 2023. doi:10.1007/s11682-022-00756-0

35. Wu X, Lin P, Yang J, Song H, Yang R, Yang J. Dysfunction of the cingulo-opercular network in first-episode medication-naive patients with major depressive disorder. J Affect Disord. 2016;200:275–283. doi:10.1016/j.jad.2016.04.046

36. Chen MH, Li CT, Lin WC, et al. Persistent antidepressant effect of low-dose ketamine and activation in the supplementary motor area and anterior cingulate cortex in treatment-resistant depression: A randomized control study. J Affect Disord. 2018;225:709–714. doi:10.1016/j.jad.2017.09.008

37. Godlewska BR, Browning M, Norbury R, Igoumenou A, Cowen PJ, Harmer CJ. Predicting Treatment Response in Depression: The Role of Anterior Cingulate Cortex. Int J Neuropsychopharmacol. 2018;21(11):988–996. doi:10.1093/ijnp/pyy069

38. Karim HT, Wang M, Andreescu C, et al. Acute trajectories of neural activation predict remission to pharmacotherapy in late-life depression. NeuroImage Clin. 2018;19:831–839. doi:10.1016/j.nicl.2018.06.006

39. Martens MAG, Filippini N, Harmer CJ, Godlewska BR. Resting state functional connectivity patterns as biomarkers of treatment response to escitalopram in patients with major depressive disorder. Psychopharmacology (Berl). 2022;239(11):3447–3460. doi:10.1007/s00213-021-05915-7

40. Li K, Lu X, Xiao C, et al. Aberrant Resting-State Functional Connectivity in MDD and the Antidepressant Treatment Effect—A 6-Month Follow-Up Study. Brain Sci. 2023;13(5):705. doi:10.3390/brainsci13050705

41. Lu F, Cui Q, Huang X, et al. Anomalous intrinsic connectivity within and between visual and auditory networks in major depressive disorder. Prog Neuropsychopharmacol Biol Psychiatry. 2020;100:109889. doi:10.1016/j.pnpbp.2020.109889

42. Dichter GS, Gibbs D, Smoski MJ. A systematic review of relations between resting-state functional-MRI and treatment response in major depressive disorder. J Affect Disord. 2015;172:8–17. doi:10.1016/j.jad.2014.09.028

43. Goldstein-Piekarski AN, Staveland BR, Ball TM, Yesavage J, Korgaonkar MS, Williams LM. Intrinsic functional connectivity predicts remission on antidepressants: a randomized controlled trial to identify clinically applicable imaging biomarkers. Transl Psychiatry. 2018;8(1):1–11. doi:10.1038/s41398-018-0100-3

44. Dunlop K, Talishinsky A, Liston C. Intrinsic Brain Network Biomarkers of Antidepressant Response: a Review. Curr Psychiatry Rep. 2019;21(9):87. doi:10.1007/s11920-019-1072-6

45. Kemp AH, Gordon E, Rush AJ, Williams LM. Improving the Prediction of Treatment Response in Depression: Integration of Clinical, Cognitive, Psychophysiological, Neuroimaging, and Genetic Measures. CNS Spectr. 2008;13(12):1066–1086. doi:10.1017/S1092852900017120

46. Pizzagalli DA. Frontocingulate Dysfunction in Depression: Toward Biomarkers of Treatment Response. Neuropsychopharmacology. 2011;36(1):183–206. doi:10.1038/npp.2010.166

47. Liston C, Chen AC, Zebley BD, et al. Default Mode Network Mechanisms of Transcranial Magnetic Stimulation in Depression. Biol Psychiatry. 2014;76(7):517–526. doi:10.1016/j.biopsych.2014.01.023

48. Posner J, Hellerstein DJ, Gat I, et al. Antidepressants Normalize the Default Mode Network in Patients With Dysthymia. JAMA Psychiatry. 2013;70(4):373–382. doi:10.1001/jamapsychiatry.2013.455

49. Chen CH, Suckling J, Ooi C, et al. Functional Coupling of the Amygdala in Depressed Patients Treated with Antidepressant Medication. Neuropsychopharmacology. 2008;33(8):1909–1918. doi:10.1038/sj.npp.1301593

50. Liu H, Wang C, Lan X, et al. Functional connectivity of the amygdala and the antidepressant and antisuicidal effects of repeated ketamine infusions in major depressive disorder. Front Neurosci. 2023;17. Accessed August 8, 2023. https://www.frontiersin.org/articles/10.3389/fnins.2023.1123797

51. Nakamura T, Tomita M, Horikawa N, et al. Functional connectivity between the amygdala and subgenual cingulate gyrus predicts the antidepressant effects of ketamine in patients with treatment-resistant depression. Neuropsychopharmacol Rep. 2021;41(2):168–178. doi:10.1002/npr2.12165

52. Salomons TV, Dunlop K, Kennedy SH, et al. Resting-State Cortico-Thalamic-Striatal Connectivity Predicts Response to Dorsomedial Prefrontal rTMS in Major Depressive Disorder. Neuropsychopharmacology. 2014;39(2):488–498. doi:10.1038/npp.2013.222

53. Alexopoulos GS, Hoptman MJ, Kanellopoulos D, Murphy CF, Lim KO, Gunning FM. Functional connectivity in the cognitive control network and the default mode network in late-life depression. J Affect Disord. 2012;139(1):56–65. doi:10.1016/j.jad.2011.12.002

