## Supplemental materials for "A common symptom geometry of mood improvement under sertraline and placebo associated with distinct neural patterns"

### Supplementary methods

#### Dimension reduction of symptom improvement

In step 1, we first performed data dimension reduction using the PCA of 73 items, as proposed and validated in our previous work.[1,2] Each item was first scaled to have unit variance across patients before running the PCA. PCA was performed for subgroups defined by treatment or clinical responses (during stage 1: sertraline, placebo, responders to sertraline, non-responders to sertraline, responders to placebo, non-responders to placebo; during stage 2: switched from sertraline to bupropion, switched from placebo to sertraline, continuing sertraline, continuing placebo; all participants during stage 1, all participants during stage 2). Importantly, in the present work, we focused on capturing the principal axes of symptom improvement between two time-points, by performing PCA on the difference of each behavioral measure (i.e. item) before and after treatment. This approach provides an overview of improvement at the item level, based on the geometry of each PC loadings. In step 2, the reproducibility of the estimated symptom PCs was evaluated using a permutation-based cross validation and null distribution based significant testing of each PC. First, significance of the derived principal components (PCs) was computed via permutation testing. For each permutation, patient order was randomly shuffled for each symptom variable before re-computing PCA. This permutation was repeated 10,000 times to establish the null model. PCs which accounted for a proportion of variance that exceeded chance ($p<0.05$ across all 10,000 permutations) were retained for further analysis. PCs are ordered as a function of the behavioral variance that they account for. Second, we performed split-half permutations (1000 times) to test for reliability of the derived PCs. PC reliability was assessed through split-half correlation: only PCs with loadings correlating above 50% across 1000 permutations were considered reliable. In step 3, in order to identify whether symptom improvement shared geometry across groups, the similarity of the geometries of estimated PCs was evaluated between two groups in each comparison, using the Pearson's correlation of the loadings of 73 behavioral measures on each PC.

**Preprocessing pipeline**

Neuroimaging data were preprocessed using the Human Connectome Project (HCP) minimal preprocessing pipeline,[3] adapted for compatibility with "legacy" data, which are now featured as a standard option in the HCP pipelines provided by our team (https://github.com/Washington-University/HCPpipelines/pull/156). These modifications to the HCP pipelines were necessary as the EMBARC data did not include a standard field map and did not incorporate a T2w high-resolution image or field maps. The adaptations for single-band BOLD acquisition have previously been described in detail.[4] The adapted HCP pipeline included the following steps: (1) The T1- weighted images were corrected for bias-field distortions and warped to the standard MNI-152 brain template through a combination of linear and nonlinear transformations using the FMRIB Software Library (FSL) linear image registration tool (FLIRT) and non-linear image registration tool (FNIRT).[5] (2) FreeSurfer's recon-all pipeline was used to compute brain-wide segmentation of gray and white matter to produce individual cortical and subcortical anatomical segmentation.[6] After completing the recon-all pipeline, all T1-weighted images were subjected to qualitative (visual) quality control (QC) by at least two independent trained researchers. (3) Cortical surface models were generated for pial and white matter boundaries as well as segmentation masks for each subcortical gray matter voxel. Using the pial and white matter surface boundaries, a "cortical ribbon" was defined along with corresponding subcortical voxels, which were combined to generate the Connectivity Informatics Technology Initiative (CIFTI) volume/surface "gray-ordinate" space for each individual subject, which drastically reduces file management for combined surface and volume analyses and visualization and establishes a combined cortical surface and subcortical volume coordinate system.[3] (4) The cortical surfaces were then registered to the group average HCP atlas using surface-based registration based on cortical landmark features, whereas the subcortical "volume" component of the image was brought into group atlas alignment via nonlinear registration.[3] (5) The BOLD data were motion corrected and aligned to the middle frame of every run via FLIRT. In turn, a liberal brain mask was applied to exclude signal from non-brain tissue. After initial processing in Neuroimaging Informatics Technology Initiative (NIFTI) volume space, BOLD data were converted to the CIFTI gray matter matrix by sampling from the anatomically defined gray matter cortical ribbon, whereas the subcortical voxels were isolated using subject-specific FreeSurfer segmentation. The subcortical volume component of the BOLD data was then aligned to the group atlas as part of the NIFTI processing in a single transform step that concatenates all of the transform matrices for each prior processing step (i.e., motion correction, registration, distortion correction). This produced a single nonlinear transformation to minimize interpolation cost. In turn, the cortical surface component of the CIFTI file was aligned to the HCP atlas using surface-based nonlinear deformation based on sulcal features. Following these "minimal" HCP preprocessing steps, a high-pass filter (0.008 Hz) was applied to the BOLD time series to remove low temporal frequencies and scanner drift. QuNex tools were then used to compute the signal in the ventricles, deep white matter, and across all gray matter voxels as proxy of global signal regression (GSR) to address spatially pervasive sources of artifacts.[7] These time series were modeled as nuisance variables and were regressed out of the gray matter voxels. Subsequent analyses used the residual BOLD time series following these denoising steps. Of note, we calculated SNR for each participant by obtaining the mean signal and SD for a given slice across the BOLD run, while excluding all non-brain voxels across all frames. In addition, we implemented "movement scrubbing," as recommended by Power.[8] Movement scrubbing refers to the practice of removing BOLD volumes that have been flagged for high motion to minimize movement artifacts. Specifically, all frames with possible movement-induced artifactual fluctuations in intensity were identified via two criteria: (1) frames displacement (the sum of the displacement across all six rigid body movement correction parameters) exceeding 0.5 mm (assuming 50 mm cortical sphere radius); and (2) the normalized root mean square (RMS) (calulated as the RMS of differences in intensity between the current and preceding frame, computed across all voxels and divided by the mean intensity) exceeding 1.6 times the median. The frames flagged by either criterion were marked for exclusion (logical or), as well as the one preceding and following the flagged frame. Subjects with >50% frames flagged were excluded from further analyses.

**Global Brain Connectivity**

We computed individual parcellated neural global brain connectivity (GBC) maps[9] [15] using a whole-brain functional network parcellation to reduce the dimensionality of the neural feature space [34]. All data was parcellated prior to conducting functional connectivity analysis as this gave the best trade-off between the sample size needed to resolve multivariate neurobehavioral solutions and the size of the feature space. For each participant, we first computed the mean BOLD signal within each parcel. Then, the individualized resting-state functional connectivity (FC) matrix was calculated by computing the Pearson's correlation between every parcel in the brain with all other parcels. A Fisher's r-to-Z transform was then applied. GBC was calculated by computing every parcel's mean FC strength with all other parcels (i.e. the mean, per row, across all columns of the FC matrix) as follows:[59]

$$GBC(x) = \frac{1}{N}\sum_{y=1}^{N} r_{xy}$$

- where $GBC(x)$ denotes the GBC value at parcel $x$;
- where $N$ denotes the total number of parcels;
- where X denotes the sum from $y$ =1 to $y = N$;
- where $r_{xy}$ denotes the correlation between the time-series of parcels $x$ and $y$;

GBC is a data-driven summary measure of connectedness that is unbiased with regards to the location of a possible alteration in connectivity[10] and is therefore a principled way for reducing the number of neural features while assessing neural variation across the entire brain.

### Supplementary results

#### Analysis of global clinical scores

In this section, we describe global clinical scores for the HRSD, ASRM, CHRT and CASTS at different time-points (baseline, 8 weeks, 16 weeks) or their evolution during stage 1 and 2 (for stage 1: baseline minus 8 weeks, and for stage 2: 8 weeks minus 16 weeks).

*Stage 1*

Patients in the placebo and the sertraline groups were similar at baseline (see Table 1), as can be observed by superimposing their symptomatology item by item (see Figure 2A). After 8 weeks, the proportion of responders and non-responders according to the CGI did not significantly differ between the placebo and the sertraline groups (placebo 39.4% vs. sertraline 51.6%, $\chi^2 = 2.4$, $p = 0.12$). When computing the difference between 8 weeks and baseline mean scores, there was a significant difference of improvement between the placebo and the sertraline groups for the CHRT scores (CHRT propensity score improvement: sertraline: mean: 11.3 (SD: 8.73) vs. placebo: 7.8 (9.53), $t_{190} = 2.63$, $p = 0.009$; CHRT risk score improvement: sertraline: 2.30 (2.49) vs. placebo: 1.12 (2.75), $t_{190} = 3.12$, $p = 0.002$) but not for the other scales (HRSD: $t_{189} = 1.20$, $p = 0.23$; ASRM: $t_{180} = -1.48$, $p = 0.14$ and CASTS: $t_{190} = 1.74$, $p = 0.08$).

When splitting the participants in each treatment group according to the clinical response (responders vs. non-responders according to the CGI at 8 weeks), we observed significant differences in symptom improvement for all scales scores (sertraline group: all $p < 0.007$; placebo group: all $p < 0.02$, see Figure 2F). However, there was no significant difference in baseline demographic or scales scores between responders and non-responders (age, gender, ethnicity, education, MDD severity, MDD chronicity, HRSD, ASMR, CHRT propensity, CHRT risk, CAST, all $p > 0.08$ across subjects and within each treatment group), suggesting that baseline symptoms are not predictive of subsequent clinical response.

*Stage 2*

Between week 8 and week 16, participants' medication depended on clinical response during stage 1: non-responders to sertraline were switched to bupropion, non-responders to placebo were switched to sertraline and responders kept their treatment the same (see Figure 1).

At the end of stage 1, non-responders to sertraline and placebo differed in their global CHRT risk score (sertraline switch to bupropion: 3.79 (2.35) vs. placebo switch to sertraline: 5.06 (2.54), $t_{75} = 2.34$, $p = 0.022$). During stage 2, they improved differently for their CHRT propensity score (sertraline (previously placebo): 8.16 (10.0) vs. bupropion (previously sertraline): 3.47 (6.1), $t_{80} = 2.65$, $p = 0.010$), CHRT risk score (sertraline: 2.0 (2.27) vs. bupropion: -0.1 (2.28), $t_{71} = 4.01$, $p = 0.0002$), and CAST scores (sertraline: 5.76 (9.27) vs. bupropion: 1.88 (7.72), $t_{78} = 2.07$, $p = 0.04$).

Responders to sertraline and placebo did not significantly differ in their scales scores at the end of stage 1 (all $p > 0.1$) and did not significantly change during stage 2 (all $p > 0.06$), nor differ in their improvements (all $p > 0.05$).

**Shared PC geometry**

Eighteen items had positive loadings above the 3rd quartile for the common PC1 of phase 1. Eight items of the HRSD: depressed mood, work and activities, worthlessness, hopelessness, helplessness, concentration, social withdrawal, fatigability; 5 items of the CHRT: I can do nothing right, everything I do turns out wrong, things will never get better, I have thoughts about how I might kill myself, no future; 4 items of the CAST: Lately everything seems to be annoying to me, I find people get on my nerves easily, I wish to be alone, anxiety. One item of the CAST had a negative loading under the 3rd quartile across all PC1: feeling good (see supplementary Figure).

### Supplementary Discussion

#### A better characterization of improvement

Mood assessment is a critical issue in psychiatry, notably because it has an impact on therapeutic strategies. In the current study, some patients were considered as non-responders and switched to another treatment while having high PC1 scores or, on the contrary, considered as responders and kept the same treatment, while having low PC1 scores. The discrepancy between classical assessment and PCA results is certainly due to the inclusion of other dimensions than pure depressive symptoms, notably suicidal risk and anxiety scores, which account for a significant part of symptom variation, especially in the sertraline arm. Indeed, most previous studies on the EMBARC dataset used the Hamilton Rating Scale for Depression (HRSD) to measure clinical response and found no superiority of sertraline over placebo,[11–16] suggesting that PC based on many items is more sensitive to subtle effects of antidepressants. How best to measure clinical improvement and which symptomatic dimensions should be included is a thorny question. Many clinical scales have been validated and are commonly used in research as well as in clinical practice.[17–22] They have strength and weakness, depending on the clinical severity and subtypes of depression that should be assessed,[23] and have therefore different fields of application.[24,25] Importantly, scales general scores can be biased by the number of items dedicated to certain domains. For instance, the Hamilton Scale includes many somatic symptoms, leading to an overestimate of antidepressant efficacy for sedative molecules and an underestimate for drugs associated with somatic side effects.[23,26] Similarly, whether patients or clinician assessments have higher sensitivity and specificity is a matter of debate.[27,28] Furthermore, global evaluation such as the CGI may be subject to rater bias.[29] All those limitations have been extensively discussed elsewhere.[30,31] With our approach, we provide a proof-of-concept that some caveats can be circumvented by an exhaustive inclusion of multiple scales items and their subsequent selection by data-driven reduction.

**Sertraline versus placebo effect**

In this study improvement geometry is widely shared between sertraline and placebo. However, our results suggest that improvement mechanisms strongly differ between the two groups. First, improvement amplitude was higher in the sertraline group, suggesting that pharmacological action amplifies placebo response. This result is in line with a previous study which found that patients under antidepressant and placebo had a similar time-course of clinical improvement but different response amplitude.[32] Second, some neural and clinical factor were specifically predictive of sertraline efficacy, suggesting that pharmacological improvement relies on more reproducible neurobehavioral features with lower heterogeneity than placebo response. Additionally, in the sertraline group, brain patterns predicting response differed according to the variable used to measure improvement (CGI versus PC1), whereas it was not the case in the placebo group. This result has two implications. First, it highlights that CGI and PC1 are more different in the sertraline group than in the placebo group. This is indeed what shows the distribution of PC1 scores: there is more discrepancy between response status and PC1 scores for sertraline than for placebo (especially among non-responders). Interestingly, sertraline improves significantly more the suicidal/anxiety dimensions (included in PC1) than placebo, suggesting that CGI could be less sensitive to those dimensions. Second, these results suggest that sertraline has different pharmacological mechanisms of action on depression and suicidal/anxiety. It could be argued that depression improvement is solely due to placebo effect, or at least that there is a strong overlap between those two.[33] However and crucially, even for response measured with CGI, sertraline and placebo maps are very dissimilar, indicating that sertraline efficacy measured by response is not reducible to a placebo effect. Therefore, pharmacological improvement seems to rely on reproducible neurobehavioral features, targeting in specific ways, depression, on the one hand, and anxiety/suicidal neural circuits, on the other hand. By contrast, placebo effect could be noisier and less differentiated in terms of brain circuits since placebo clinical response was generally less predicted by brain (and clinical) patterns compared to sertraline response. Overall, enriching improvement measures with clinical features other than depression reveals circuits that seem to be specific to the pharmacological action and go beyond placebo response.

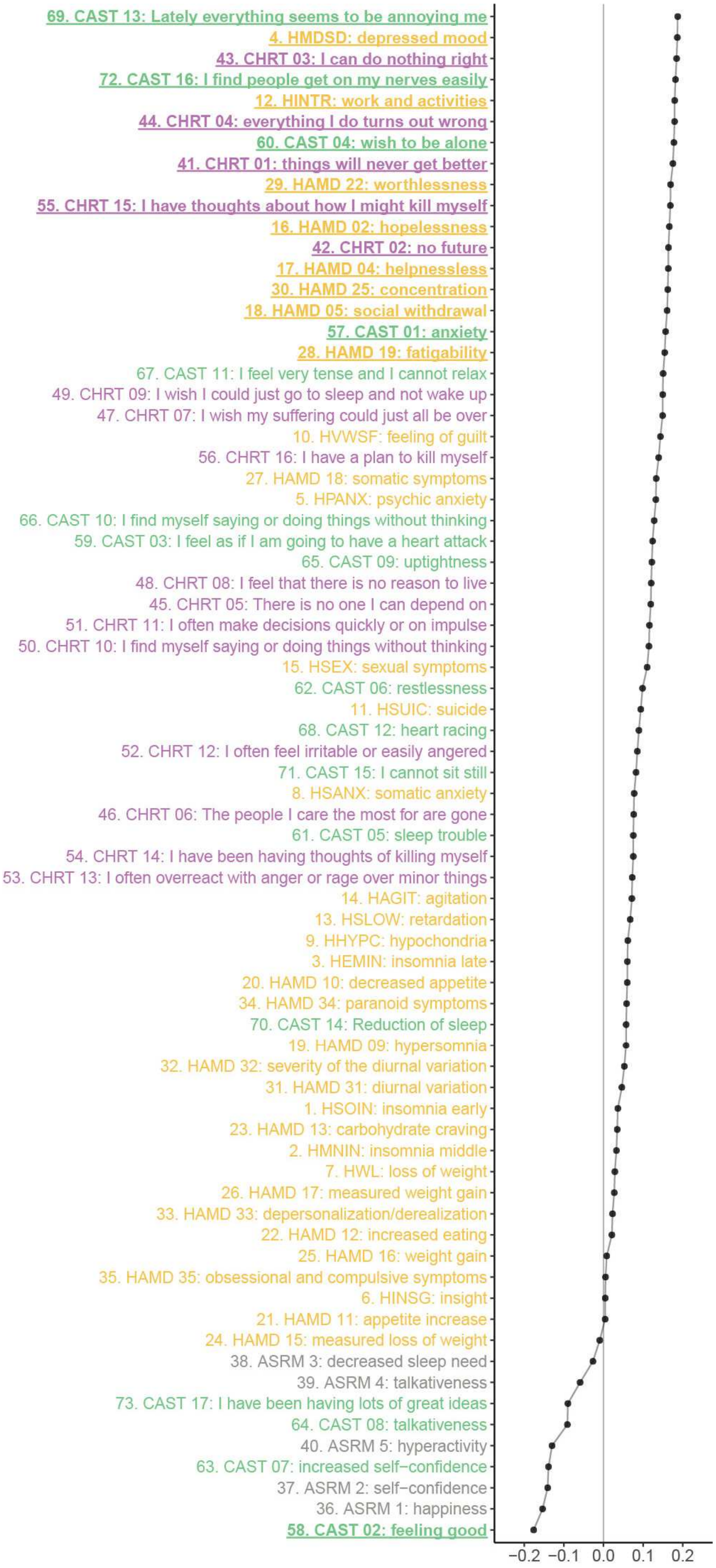

69. CAST 13: Lately everything seems to be annoying me
4. HMDSD: depressed mood
43. CHRT 03: I can do nothing right
72. CAST 16: I find people get on my nerves easily
12. HINTR: work and activities
44. CHRT 04: everything I do turns out wrong
60. CAST 04: wish to be alone
41. CHRT 01: things will never get better
29. HAMD 22: worthlessness
55. CHRT 15: I have thoughts about how I might kill myself
16. HAMD 02: hopelessness
42. CHRT 02: no future
17. HAMD 04: helpnessless
30. HAMD 25: concentration
18. HAMD 05: social withdrawal
57. CAST 01: anxiety
28. HAMD 19: fatigability
67. CAST 11: I feel very tense and I cannot relax
49. CHRT 09: I wish I could just go to sleep and not wake up
47. CHRT 07: I wish my suffering could just all be over
10. HVWSF: feeling of guilt
56. CHRT 16: I have a plan to kill myself
27. HAMD 18: somatic symptoms
5. HPANX: psychic anxiety
66. CAST 10: I find myself saying or doing things without thinking
59. CAST 03: I feel as if I am going to have a heart attack
65. CAST 09: uptightness
48. CHRT 08: I feel that there is no reason to live
45. CHRT 05: There is no one I can depend on
51. CHRT 11: I often make decisions quickly or on impulse
50. CHRT 10: I find myself saying or doing things without thinking
15. HSEX: sexual symptoms
62. CAST 06: restlessness
11. HSUIC: suicide
68. CAST 12: heart racing
52. CHRT 12: I often feel irritable or easily angered
71. CAST 15: I cannot sit still
8. HSANX: somatic anxiety
46. CHRT 06: The people I care the most for are gone
61. CAST 05: sleep trouble
54. CHRT 14: I have been having thoughts of killing myself
53. CHRT 13: I often overreact with anger or rage over minor things
14. HAGIT: agitation
13. HSLOW: retardation
9. HHYPC: hypochondria
3. HEMIN: insomnia late
20. HAMD 10: decreased appetite
34. HAMD 34: paranoid symptoms
70. CAST 14: Reduction of sleep
19. HAMD 09: hypersomnia
32. HAMD 32: severity of the diurnal variation
31. HAMD 31: diurnal variation
1. HSOIN: insomnia early
23. HAMD 13: carbohydrate craving
2. HMNIN: insomnia middle
7. HWL: loss of weight
26. HAMD 17: measured weight gain
33. HAMD 33: depersonalization/derealization
22. HAMD 12: increased eating
25. HAMD 16: weight gain
35. HAMD 35: obsessional and compulsive symptoms
6. HINSG: insight
21. HAMD 11: appetite increase
24. HAMD 15: measured loss of weight
38. ASRM 3: decreased sleep need
39. ASRM 4: talkativeness
73. CAST 17: I have been having lots of great ideas
64. CAST 08: talkativeness
40. ASRM 5: hyperactivity
63. CAST 07: increased self−confidence
37. ASRM 2: self−confidence
36. ASRM 1: happiness
58. CAST 02: feeling good
−0.2
−0.1
0.0
0.1
0.2

**Supplementary Figure 1. Loadings of common PC1 during phase 1.** The items above the 3rd quartile in absolute value are underlined

### References


1. Moujaes F, Ji JL, Rahmati M, et al. *Ketamine Induces Multiple Individually Distinct Whole-Brain Functional Connectivity Signatures*. Neuroscience; 2022. doi:10.1101/2022.11.01.514692

2. Ji JL, Diehl C, Schleifer C, et al. Schizophrenia Exhibits Bi-directional Brain-Wide Alterations in Cortico-Striato-Cerebellar Circuits. *Cereb Cortex N Y N 1991*. 2019;29(11):4463-4487. doi:10.1093/cercor/bhy306

3. Glasser MF, Sotiropoulos SN, Wilson JA, et al. The minimal preprocessing pipelines for the Human Connectome Project. *NeuroImage*. 2013;80:105-124. doi:10.1016/j.neuroimage.2013.04.127

4. Ji JL, Spronk M, Kulkarni K, Repovš G, Anticevic A, Cole MW. Mapping the human brain's cortical-subcortical functional network organization. *NeuroImage*. 2019;185:35-57. doi:10.1016/j.neuroimage.2018.10.006

5. Jenkinson M, Bannister P, Brady M, Smith S. Improved optimization for the robust and accurate linear registration and motion correction of brain images. *NeuroImage*. 2002;17(2):825-841. doi:10.1016/s1053-8119(02)91132-8

6. Reuter M, Schmansky NJ, Rosas HD, Fischl B. Within-subject template estimation for unbiased longitudinal image analysis. *NeuroImage*. 2012;61(4):1402-1418. doi:10.1016/j.neuroimage.2012.02.084

7. Power JD, Plitt M, Laumann TO, Martin A. Sources and implications of whole-brain fMRI signals in humans. *NeuroImage*. 2017;146:609-625. doi:10.1016/j.neuroimage.2016.09.038

8. Power JD, Barnes KA, Snyder AZ, Schlaggar BL, Petersen SE. Spurious but systematic correlations in functional connectivity MRI networks arise from subject motion. *NeuroImage*. 2012;59(3):2142-2154. doi:10.1016/j.neuroimage.2011.10.018

9. Cole MW, Pathak S, Schneider W. Identifying the brain's most globally connected regions. *NeuroImage*. 2010;49(4):3132-3148. doi:10.1016/j.neuroimage.2009.11.001

10. Cole MW, Yang GJ, Murray JD, Repovš G, Anticevic A. Functional connectivity change as shared signal dynamics. *J Neurosci Methods*. 2016;259:22-39. doi:10.1016/j.jneumeth.2015.11.011

11. Webb CA, Trivedi MH, Cohen ZD, et al. Personalized prediction of antidepressant v. placebo response: evidence from the EMBARC study. *Psychol Med*. 2019;49(7):1118-1127. doi:10.1017/S0033291718001708

12. Chin Fatt CR, Jha MK, Cooper CM, et al. Effect of Intrinsic Patterns of Functional Brain Connectivity in Moderating Antidepressant Treatment Response in Major Depression. *Am J Psychiatry*. 2020;177(2):143-154. doi:10.1176/appi.ajp.2019.18070870

13. Fan S, Nemati S, Akiki TJ, et al. Pretreatment Brain Connectome Fingerprint Predicts Treatment Response in Major Depressive Disorder. *Chronic Stress*. 2020;4:2470547020984726. doi:10.1177/2470547020984726

14. Whitton AE, Webb CA, Dillon DG, et al. Pretreatment Rostral Anterior Cingulate Cortex Connectivity With Salience Network Predicts Depression Recovery: Findings From the EMBARC Randomized Clinical Trial. *Biol Psychiatry*. 2019;85(10):872-880. doi:10.1016/j.biopsych.2018.12.007

15. Cooper CM, Chin Fatt CR, Jha M, et al. Cerebral Blood Perfusion Predicts Response to Sertraline versus Placebo for Major Depressive Disorder in the EMBARC Trial. *EClinicalMedicine*. 2019;10:32-41. doi:10.1016/j.eclinm.2019.04.007

16. Pizzagalli DA, Webb CA, Dillon DG, et al. Pretreatment Rostral Anterior Cingulate Cortex Theta Activity in Relation to Symptom Improvement in Depression: A Randomized Clinical Trial. *JAMA Psychiatry*. 2018;75(6):547-554. doi:10.1001/jamapsychiatry.2018.0252

17. Montgomery SA, Åsberg M. A New Depression Scale Designed to be Sensitive to Change. *Br J Psychiatry*. 1979;134(4):382-389. doi:10.1192/bjp.134.4.382

18. Hamilton M. A rating scale for depression. *J Neurol Neurosurg Psychiatry*. 1960;23(1):56-62. doi:10.1136/jnnp.23.1.56

19. Trajković G, Starčević V, Latas M, et al. Reliability of the Hamilton Rating Scale for Depression: A meta-analysis over a period of 49years. *Psychiatry Res*. 2011;189(1):1-9. doi:10.1016/j.psychres.2010.12.007

20. Rush AJ, Trivedi MH, Ibrahim HM, et al. The 16-Item quick inventory of depressive symptomatology (QIDS), clinician rating (QIDS-C), and self-report (QIDS-SR): a psychometric evaluation in patients with chronic major depression. *Biol Psychiatry*. 2003;54(5):573-583. doi:10.1016/S0006-3223(02)01866-8

21. Beck AT, Ward CH, Mendelson M, Mock J, Erbaugh J. An Inventory for Measuring Depression. *Arch Gen Psychiatry*. 1961;4(6):561-571. doi:10.1001/archpsyc.1961.01710120031004

22. Trull TJ, Ebner-Priemer UW. Using Experience Sampling Methods/Ecological Momentary Assessment (ESM/EMA) in Clinical Assessment and Clinical Research: Introduction to the Special Section. *Psychol Assess*. 2009;21(4):457-462. doi:10.1037/a0017653

23. Möller HJ. Methodological aspects in the assessment of severity of depression by the Hamilton Depression Scale. *Eur Arch Psychiatry Clin Neurosci*. 2001;251 Suppl 2:II13-20. doi:10.1007/BF03035121

24. Furukawa TA. Assessment of mood: Guides for clinicians. *J Psychosom Res*. 2010;68(6):581-589. doi:10.1016/j.jpsychores.2009.05.003

25. Nezu AM, McClure KS, Nezu CM. The Assessment of Depression. In: *Treating Depression*. John Wiley & Sons, Ltd; 2015:24-51. doi:10.1002/9781119114482.ch2

26. Maier W, Philipp M. Improving the Assessment of Severity of Depressive States: A Reduction of the Hamilton Depression Scale. *Pharmacopsychiatry*. 1985;18(01):114-115. doi:10.1055/s-2007-1017335

27. Chevance A, Gourion D, Hoertel N, et al. Ensuring mental health care during the SARS-CoV-2 epidemic in France: A narrative review. *L'Encéphale*. 2020;46(3):193-201. doi:10.1016/j.encep.2020.04.005

28. Bailey J, Coppen A. A Comparison Between the Hamilton Rating Scale and the Beck Inventory in the Measurement of Depression. *Br J Psychiatry*. 1976;128(5):486-489. doi:10.1192/bjp.128.5.486

29. Petkova E, Quitkin FM, McGrath PJ, Stewart JW, Klein DF. A Method to Quantify Rater Bias in Antidepressant Trials. *Neuropsychopharmacology*. 2000;22(6):559-565. doi:10.1016/S0893-133X(99)00154-2

30. Demyttenaere K, De Fruyt J. Getting What You Ask For: On the Selectivity of Depression Rating Scales. *Psychother Psychosom*. 2003;72(2):61-70. doi:10.1159/000068690

31. Cusin C, Yang H, Yeung A, Fava M. Rating Scales for Depression. In: Baer L, Blais MA, eds. *Handbook of Clinical Rating Scales and Assessment in Psychiatry and Mental Health*. Current Clinical Psychiatry. Humana Press; 2010:7-35. doi:10.1007/978-1-59745-387-5_2

32. Gueorguieva R, Mallinckrodt C, Krystal JH. Trajectories of depression severity in clinical trials of duloxetine: insights into antidepressant and placebo responses. *Arch Gen Psychiatry*. 2011;68(12):1227-1237. doi:10.1001/archgenpsychiatry.2011.132

33. Huneke NTM, Aslan IH, Fagan H, et al. Functional Neuroimaging Correlates of Placebo Response in Patients With Depressive or Anxiety Disorders: A Systematic Review. *Int J Neuropsychopharmacol*. 2022;25(6):433-447. doi:10.1093/ijnp/pyac009
