## Supplementary figures and images for "A common symptom geometry of mood improvement under sertraline and placebo associated with distinct neural patterns"

### Supplemental Figure 1

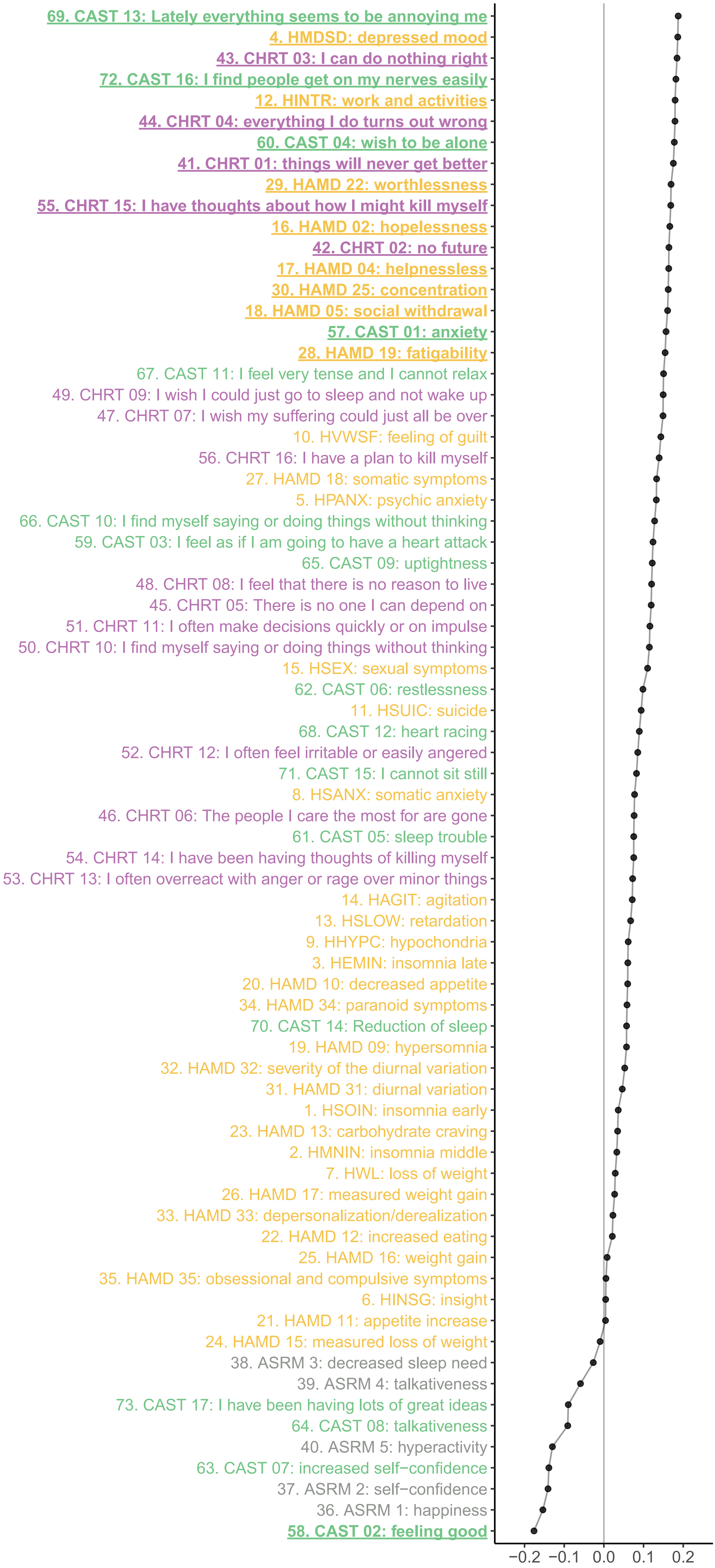
